# Causal Associations Between Imaging-derived Phenotypes and Risk of Alzheimer’s Disease and Other Neurodegenerative Disorders: A Mendelian Randomization Study

**DOI:** 10.1101/2024.09.10.24313402

**Authors:** Zhichun Chen, Jun Liu, Yong You

**Affiliations:** Department of Neurology, The Second Affiliated Hospital of Hainan Medical University, Haikou 570311, China; Department of Neurology and Institute of Neurology, Ruijin Hospital affiliated to Shanghai Jiao Tong University School of Medicine, Shanghai, 200025, China; International Center for Aging and Cancer (ICAC), Hainan Medical University; Key Laboratory of Brain Science Research & Transformation in Tropical Environment of Hainan Province

**Keywords:** Alzheimer’s disease, Imaging-derived phenotypes, Mendelian randomization, Neurodegenerative disease

## Abstract

**Background:** Accumulating observational studies have suggested associations between imaging-derived phenotypes (IDPs) and common neurodegenerative disorders, especially Alzheimer’s disease (AD). The goal of this study is to evaluate the causal associations between structural and functional IDPs and 4 neurodegenerative disorders, including AD, Parkinson’s disease (PD), Amyotrophic lateral sclerosis (ALS), and Multiple sclerosis (MS).

**Methods:** Bidirectional two-sample Mendelian randomization (MR) studies were conducted using summary statistics obtained from genome-wide association studies of 3909 IDPs from UK biobank and 4 neurodegenerative disorders.

**Results:** Forward MR analysis showed that volume of cerebral white matter in the left hemisphere was associated with increased risk of ALS (odds ratio [OR] = 1.15, 95% confidence interval [CI] = 1.09-1.22, *P* = 3.52 x 10^-6^). In reverse MR analysis, we revealed genetically determined risk of AD and MS were associated with multiple IDPs (all *P* < 1.28 x 10^-5^[0.05/3909], 9 IDPs in AD and 4 IDPs in MS). For example, genetically determined risk of AD was causally associated with reduced volume of gray matter in right ventral striatum (OR = 0.95, 95% CI = 0.93-0.97, *P* = 4.68 x 10^-7^) and lower rfMRI amplitudes in several nodes (ICA25 node 9, ICA25 node 8, and ICA100 node 11). Additionally, genetically determined risk of MS was causally associated with reduced volume in left putamen (OR = 0.97, 95% CI = 0.97-0.98, *P* = 4.47 x 10^-7^) and increased orientation dispersion index in right hippocampus (OR = 1.03, 95% CI = 1.01-1.04, *P* = 2.02 x 10^-6^).

**Conclusions:** Our study suggested plausible causal associations between risk of NDDs and brain IDPs. These findings might hold promise for identifying new disease mechanisms and developing novel preventative therapies for NDDs at the brain imaging levels.

## Background

Neurodegenerative disorders (NDDs) are characterized by the chronic degeneration of nerve cells in central or peripheral nervous system, finally leading to motor, cognitive, emotional, and autonomic disturbances. The prevalence of NDDs is increasing in part due to the extensions of lifespan, however, there is still no cure for any of these diseases. The most common NDDs include Alzheimer’s disease (AD) and Parkinson’s disease (PD), which significantly decrease life expectancy and quality of life^1, 2^.

All the NDDs exhibit characteristic imaging-derived phenotypes (IDPs) in magnetic resonance imaging (MRI) examinations, such as medial temporal lobe atrophy in AD and white matter lesions in MS. Indeed, a multitude of observational studies have been conducted to investigate the relationships between brain IDPs and NDDs. Previous studies have tried to identify potential IDPs that may affect the risk of NDDs. For example, specific brain atrophy patterns have been revealed to be associated with faster cognitive decline and highest risk of developing AD^3^. Fractional anisotropy in fornix has been demonstrated to predict the progression of mild cognitive impairment to AD in a longitudinal study^4^. Besides, specific MRI characteristics have been utilized to predict the diagnosis of multiple sclerosis (MS) in children with CNS demyelination^5^. In addition to focusing on the effects of brain IDPs on risk of NDDs, many studies have used MRI to assess the changes of brain IDPs in patients diagnosed with NDDs. For example, PD patients exhibited reduced cortical-subcortical sensorimotor connectivity compared with healthy control participants^6^. AD patients exhibited increases in cortical thickness and decreases in cortical diffusivity in early preclinical stage, but reduced cortical thickness and increased cortical diffusivity in symptomatic stage^7^. Although accumulated studies have reported potential relationships between brain IDPs and NDDs, whether brain IDPs causally affect the risk of NDDs or whether NDDs causally affect brain IDPs remain largely unknown.

Mendelian randomization (MR) is a powerful statistical method that utilizes genetic variants as instruments to infer the causal associations between exposure and outcome^8, 9^. Considering the critical role of brain IDPs in the occurrence of NDDs and NDD-associated clinical symptoms, it is important to examine whether brain IDPs are causally associated with NDDs using MR analysis. Stephen *et al*. (2021) previously published genome-wide associations of 3,935 brain IDPs, thus providing a valuable source to examine the causal associations between brain IDPs and NDDs using bidirectional MR analysis. According to previous studies, the MR analyses have demonstrated causal associations between several brain IDPs and education attainment^10^, cognitive impairment^11^, stroke^12^, and common psychiatric disorders^13, 14, 15^. In this study, we used two-sample bidirectional MR method to systematically investigate the causal associations between 3909 IDPs and 4 NDDs. Our study might provide new insights into the diagnosis and treatment of common NDDs.

## Methods

### Ethical approval

This MR study is performed based on the open-access summary statistics from previous published genome-wide association studies (GWAS) with ethical approval by corresponding ethics committee, thereby, additional ethical approval is not required.

### Study design

The overall study design is illustrated in Fig. 1. MR analyses were conducted in accordance with the STROBE-MR checklist ^16^ and Burgess *et al*.’s guidelines^17^. In forward MR analysis, we treated each brain IDP as the exposure (a total of 3909 IDPs; sample size: n = 27663 ∼ 33219)^18^ and the risk of each NDD (PD, AD, Amyotrophic lateral sclerosis [ALS], and MS; sample size: n = 115,803 ∼ 482,730) as the outcome^19, 20, 21, 22^. In reverse MR analysis, the risk of each NDD was set as the exposure and each brain IDP as the outcome.

**Fig. 1.**
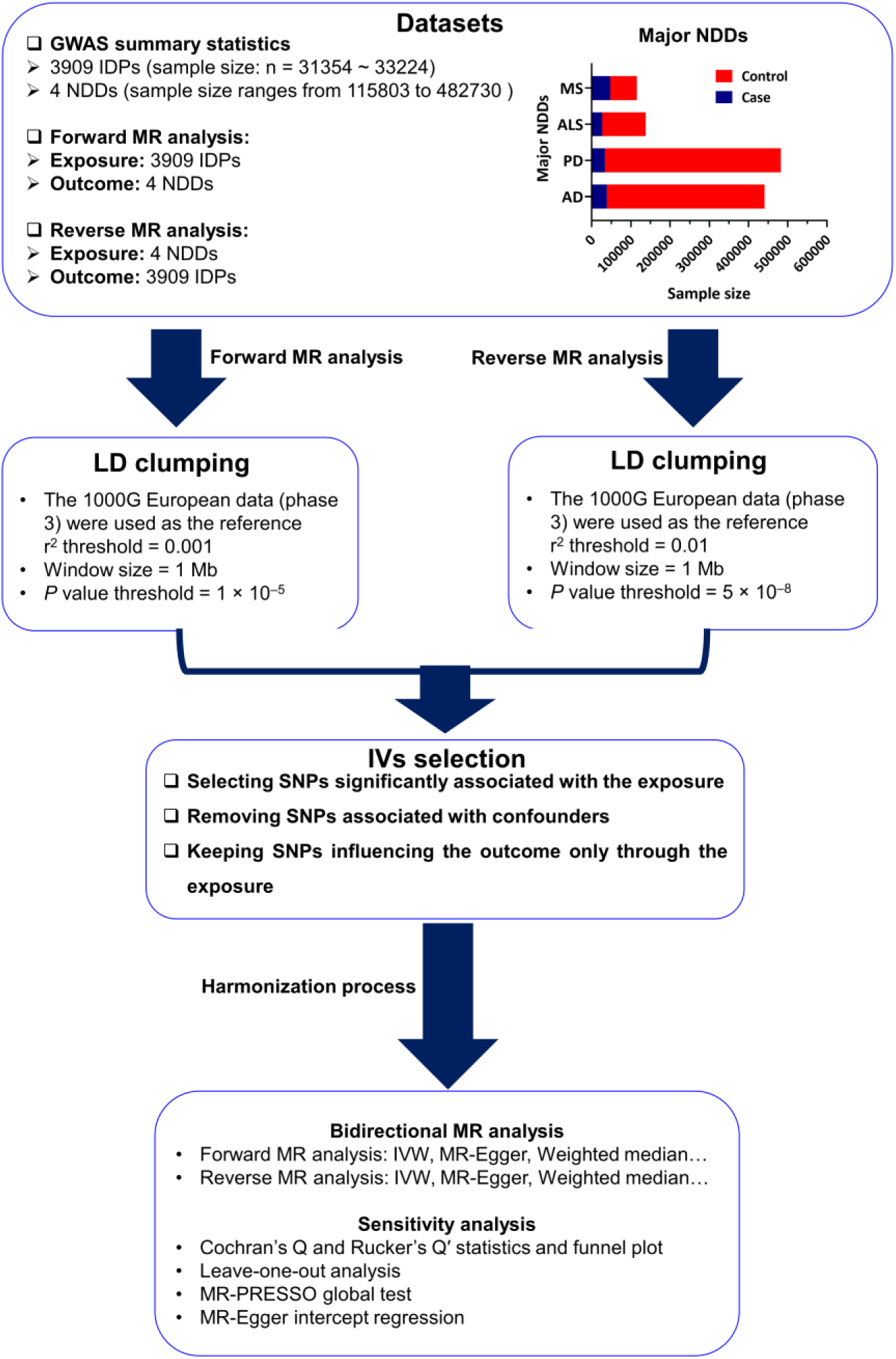
Study design of the causal inference between brain IDPs and common NDDs. Abbreviations: NDD, Neurodegenerative disorder; AD, Alzheimer’s disease; PD, Parkinson’s disease; ALS, amyotrophic lateral sclerosis; MS, multiple sclerosis; GWAS, Genome-wide association study; SNP, single nucleotide polymorphism; IV, Instrumental variable; LD, Linkage disequilibrium; MAF, minor allele frequency; MR, Mendelian randomization; IVW, Inverse variance weighted; MR-RAPS, Mendelian randomization-robust adjusted profile score; MR-PRESSO, Mendelian Randomization Pleiotropy RESidual Sum and Outlier.

### Exposure data

In forward MR analysis, the GWAS data of brain IDPs were sourced from BIG40 (https://open.win.ox.ac.uk/ukbiobank/big40/)^18^ and the IEU OpenGWAS project. This dataset comprised the genome-wide mapping of 3935 brain IDPs derived from a sample size of 33,224 individuals of European ancestry from the UK Biobank release 2020^18^. To avoid meaningless statistical analyses, we excluded several IDPs from primary 3935 IDPs, such as quality control-related IDPs. We kept nearly all brain IDPs derived from different parcellation schemes, considering that parcellation methods significantly affect the imaging properties of brain IDPs. We also included several brain IDPs that are relatively less investigated according to previous literature, including IDPs based on cortical grey-white contrast and tissue intensity. The finally selected 3909 IDPs investigated in this study were listed in Additional file 1: Table S1. They were divided into 4 categories: structural MRI IDPs, diffusion MRI (dMRI)-based IDPs, functional MRI (fMRI)-based IDPs, and susceptibility-weighted imaging (SWI)-based IDPs. In reverse MR analysis, the summary statistics of 4 NDDs, including PD (GWAS ID: ieu-b-7)^20^, AD (GWAS ID: ieu-b-2)^21^, ALS (GWAS ID: ebi-a-GCST90027163)^22^ and MS (GWAS ID: ieu-b-18)^19^, were all extracted from IEU OpenGWAS project (https://gwas.mrcieu.ac.uk/). All participants included in above original GWAS were of European ancestry. The GWAS summary statistics of 4 NDDs have been widely used in previous MR studies^23, 24^.

### Outcome data

In forward MR analysis, the GWAS summary statistics of each NDD were obtained from IEU OpenGWAS project (https://gwas.mrcieu.ac.uk/) as above described. In reverse MR analysis, the summary statistics of 3909 brain IDPs were sourced from IEU OpenGWAS project (https://gwas.mrcieu.ac.uk/).

### Selection of genetic IVs

According to the standard procedures of MR studies^8, 9, 16, 17^, each instrumental variable (IV) in this study should meet 3 assumptions as described below: (i) the IV significantly correlated with the exposure; (ii) the IV was independent of all other IVs and not associated with confounders; (iii) the IV affected the outcome only via the exposure. During the MR analysis, multiple statistical approaches such as inverse variance weighted (IVW) method and weighted median method were performed, but only IVW was selected as the main method to infer causal relationships between the exposure and outcome due to its high statistical power^13, 23^.

To increase the statistical power of the forward MR analysis, our study initially used a relaxed statistical *P*-value threshold (*P* < 1 x 10^-5^) to screen for IVs as previously described^14^. Then, the linkage disequilibrium (LD) clumping was further applied within a 10 MB window using 1000 Genomes Project Phase 3 reference panel for the European populations to identify SNPs that were independently (r^2^ < 0.001) associated with brain IDPs. In the reverse MR analysis, *P* < 5 x 10^-8^ and r^2^ < 0.01 were set to screen the IVs for each NDD.

If a particular requested SNP was not present in the outcome GWAS, then highly correlated proxy SNPs (r^2^ > 0.8) were searched as IVs. F-statistics were performed to measure the power of IVs and F-statistic < 10 indicates greater bias.

### Removing confounders

SNPs significantly associated with confounding factors, such as drinking and smoking, were excluded. Specifically, PhenoScanner V2 (http://www.phenoscanner.medschl.cam.ac.uk/) and NHGRI-EBI GWAS catalog (https://www.ebi.ac.uk/gwas/docs/file-downloads/) were used to exclude the SNPs associated with confounders.

### Two-sample MR analyses

Initially, an IVW regression method with a fixed-effects model was utilized as the primary causal inference. If the heterogeneity test is significant, a random-effects model was used during IVW regression. Complementary MR methods included MR Egger, weighted median, simple mode, and weighted mode, which might strengthen the reliability of the IVW estimates. In forward MR analysis, MR-RAPS (Robust adjusted profile score)^25^ was also used to measure the robustness of the IVW estimates because of the usage of relatively weak IVs^25^. These statistical methods were realized by the functions ‘mr_ivw’, ‘mr_egger_regression’, ‘mr_weighted_median’, ‘mr_raps’, and ‘mr_weighted_mode’ in the TwoSampleMR v0.4.26 R package. Bonferroni-corrected *P* < 0.05 (uncorrected *P* < 1.28 x 10^-5^ [0.05/3909]) was considered statistically significant. Bonferroni-corrected *P* < 0.05 within each category of IDPs was suggested to have potential significance.

### Sensitivity analysis

First, heterogeneity test using Cochran’s *Q* and Rucker’s *Q*′ statistics^26^ and funnel plot were utilized to assess the heterogeneity of IVs. Second, leave-one-out (LOO) analysis was conducted to assess whether the causal association was mainly driven by a single IV and IVs with *P* < 0.05 were regarded outliers. Third, MR-PRESSO global test (https://github.com/rondolab/MR-PRESSO/) was utilized to detect IVs having horizontal pleiotropy. Finally, an MR-Egger regression was conducted to detect the potential bias caused by directional pleiotropy. The intercept in the Egger regression indicated the existence of directional pleiotropy when the value differed from zero.

### Data availability

GWAS statistics of brain IDPs were collected from BIG40 web browser (https://open.win.ox.ac.uk/ukbiobank/big40).

### Code availability

All software packages we used in the study are publicly available, and the download links are included in the “Methods” section. The Code was available from the corresponding author upon reasonable request.

## Results

### Forward MR: the putative causal effects of IDPs on NDDs

In the forward MR analysis, we found no brain IDPs were causally associated with the risk of AD, PD, and MS (all *P* > 1.28 x 10^-5^; Additional file 1: Fig.S1-3). However, several brain IDPs might exhibit suggestive associations with above diseases (*P* < 1.00 x 10^-4^). For example, higher cortical thickness in superior medial temporal pole of left hemisphere was potentially associated with reduced risk of AD (OR = 0.68, 95%CI = 0.56-0.82, *P* = 5.96 x 10^-5^; Additional file 1: Fig. S1). Additionally, greater area in left lateral orbitofrontal cortex might be associated with a lower risk of MS (OR = 0.79, 95%CI = 0.71-0.88, *P* = 4.54 x 10^-5^; Additional file 1: Fig. S3). Importantly, we revealed the higher volume of cerebral white matter in the left hemisphere was causally associated with higher risk of ALS (odds ratio [OR] = 1.15, 95% confidence interval [CI] = 1.09-1.22, *P* = 3.52 x 10^-6^; Fig. 2), which passed the sensitivity analysis.

**Fig. 2.**
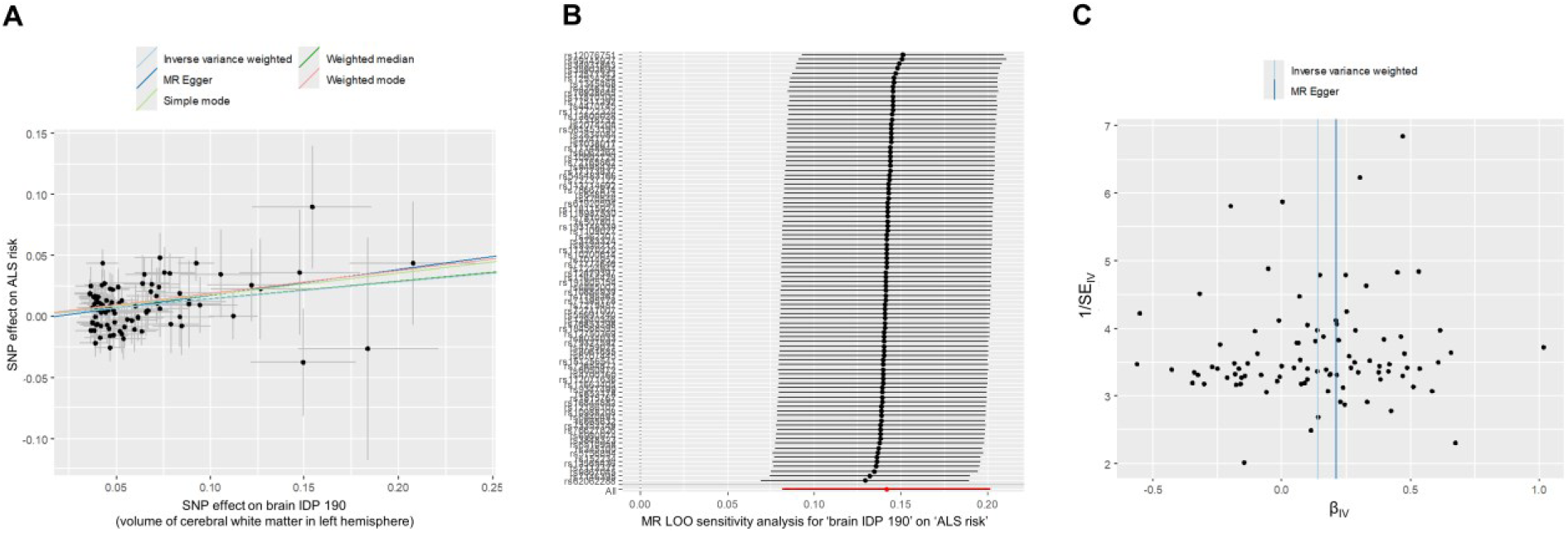
Putatively causal effects of volume of cerebral white matter in the left hemisphere on ALS risk. (A) Scatter plot for MR analysis between brain IDP 190 (volume of cerebral white matter in the left hemisphere) and ALS risk. (B) LOO sensitivity analysis for MR estimation in A. (C) Funnel plot for heterogeneity analysis in MR analysis. All statistical tests were two sided. *p* < 1.28 × 10^−5^ after Bonferroni correction was considered significant. Causal effects were estimated using five two-sample MR methods (MR-Egger, IVW, weighted median, weighted mode, and simple mode). Abbreviations: ALS, Amyotrophic lateral sclerosis; SNP, Single nucleotide polymorphism; IDP, Imaging-derived phenotype; LOO, Leave-One-Out.

### Reverse MR: the putative causal effects of NDDs on IDPs

In the reverse MR analysis, we found genetically determined risk of AD and MS were causally associated with multiple brain IDPs (all *P* < 1.28 x 10^-5^, 9 IDPs in AD and 4 IDPs in MS). Particularly, in structural MRI IDPs, genetically determined risk of AD was causally associated with reduced volume of grey matter in right ventral striatum (OR = 1.06, 95% CI = 1.04-1.08, *P* = 1.72 x 10^-9^; Table 1, Fig. 3-4, Additional file 1: Fig. S4-5) and increased mean intensity in right amygdala (OR = 1.05, 95% CI = 1.03-1.07, *P* = 4.68 x 10^-7^; Table 1, Fig. 3-4, Additional file 1: Fig. S4-5) and right Accumbens area (OR = 1.05, 95% CI = 1.03-1.07, *P* = 3.33 x 10^-7^; Table 1, Fig. 3-4, Additional file 1: Fig. S4-5). The genetically determined risk of AD was also causally associated with lower intensity-contrast in right inferior temporal gyrus (OR = 0.95, 95% CI = 0.93-0.97, *P* = 4.54 x 10^-6^; Table 1, Fig. 3-4, Additional file 1: Fig. S4-5) and right parahippocampal gyrus (OR = 0.96, 95% CI = 0.94-0.98, *P* = 1.02 x 10^-5^; Table 1, Fig. 3-4, Additional file 1: Fig. S4-5). In functional MRI IDPs, genetically determined risk of AD was causally associated with lower rfMRI amplitudes in several nodes, including ICA25 node 9, ICA25 node 8, and ICA100 node 11 (Table 1, Fig. 3-4, Additional file 1: Fig. S4-5) and increased rfMRI connectivity (ICA100 edge 681; Table 1, Fig. 3-4, Additional file 1: Fig. S4-5). For MS, genetically determined risk of MS was causally associated with reduced volume in left putamen (OR = 0.97, 95% CI = 0.97-0.98, *P* = 4.47 x 10^-7^; Table 1, Fig. 5-6, Additional file 1: Fig.S6-7) and increased orientation dispersion index in right hippocampus (OR = 1.03, 95% CI = 1.01-1.04, *P* = 2.02 x 10^-6^; Table 1, Fig. 5-6, Additional file 1: Fig. S6-7) and higher area in parahippocampal gyrus in both DKTatlas (OR = 1.02, 95% CI = 1.01-1.03, *P* = 3.21 x 10^-6^; Table 1, Fig. 5-6, Additional file 1: Fig. S6-7) and Desikan-Killiany parcellation (OR = 1.02, 95% CI = 1.01-1.03, *P* = 3.37 x 10^-6^; Table 1, Fig. 5-6, Additional file 1: Fig. S6-7). The genetically determined risk of PD and ALS were not found to be causally associated with brain IDPs (all *P* > 1.28 x 10^-5^; Additional file 1: Fig. S8-9).

**Fig. 3.**
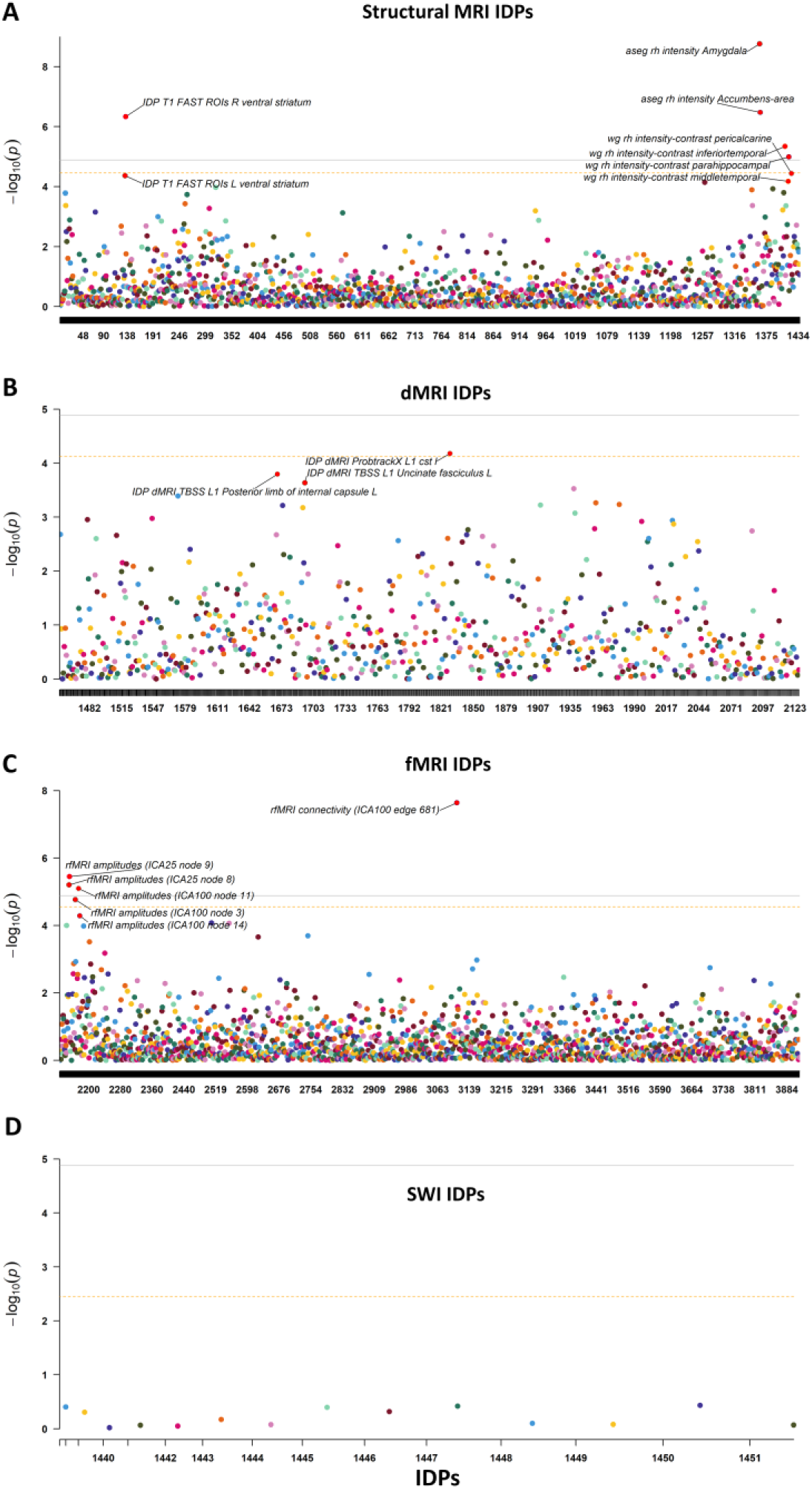
Causal associations between AD risk and brain IDPs in reverse MR analysis. (A) Causal associations between AD risk and 1433 structural MRI IDPs. (B) Causal associations between AD risk and 675 dMRI MRI IDPs. (C) Causal associations between AD risk and 1787 fMRI IDPs. (D) Causal associations between AD risk and 14 SWI IDPs. Causal effects were estimated using IVW method. Bonferroni-corrected *P* < 0.05 (uncorrected *P* < 1.28 x 10^-5^ [0.05/3909]) was considered statistically significant (Solid line). Bonferroni-corrected *P* < 0.05 within each category of IDPs was suggested to have potential significance (Dashed line). Abbreviations: IDP, Imaging-derived phenotype; MRI, Magnetic resonance imaging; SWI, Susceptibility-weighted imaging.

**Fig. 4.**
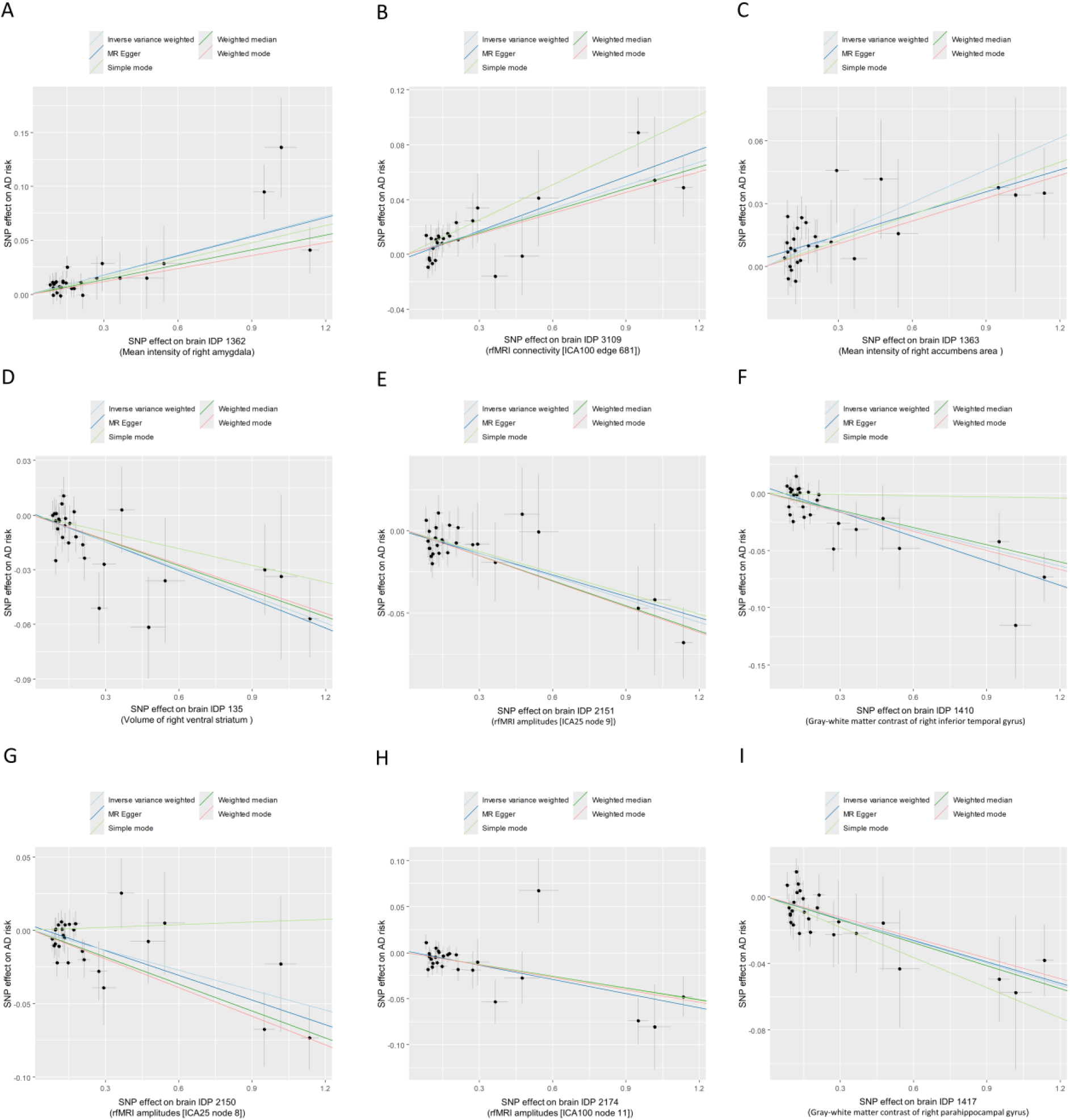
Putatively causal effects of AD risk on brain IDPs. (A-I) Causal effects of AD risk on 9 brain IDPs: IDP 1362(A, Mean intensity of right amygdala), IDP 3109 (B, rfMRI connectivity [ICA100 edge 681]), IDP 1363 (C, Mean intensity of right accumbens area), IDP 135 (D, Volume of right ventral striatum), IDP 2151(E, rfMRI amplitudes [ICA25 node 9]), IDP 1410 (F, Gray-white matter contrast of right inferior temporal gyrus), IDP 2150 (G, rfMRI amplitudes [ICA25 node 8]), IDP 2174 (H, rfMRI amplitudes [ICA100 node 11]), and IDP 1417 (I, Gray-white matter contrast of right parahippocampal gyrus). Causal effects were estimated using five two-sample MR methods (MR-Egger, IVW, weighted median, weighted mode, and simple mode). Bonferroni-corrected *P* < 0.05 (uncorrected *P* < 1.28 x 10^-5^ [0.05/3909]) was considered statistically significant. Abbreviations: AD, Alzheimer’s disease; IDP, Imaging-derived phenotype; SNP, Single nucleotide polymorphism; rfMRI, resting-state functional magnetic resonance imaging.

**Fig. 5.**
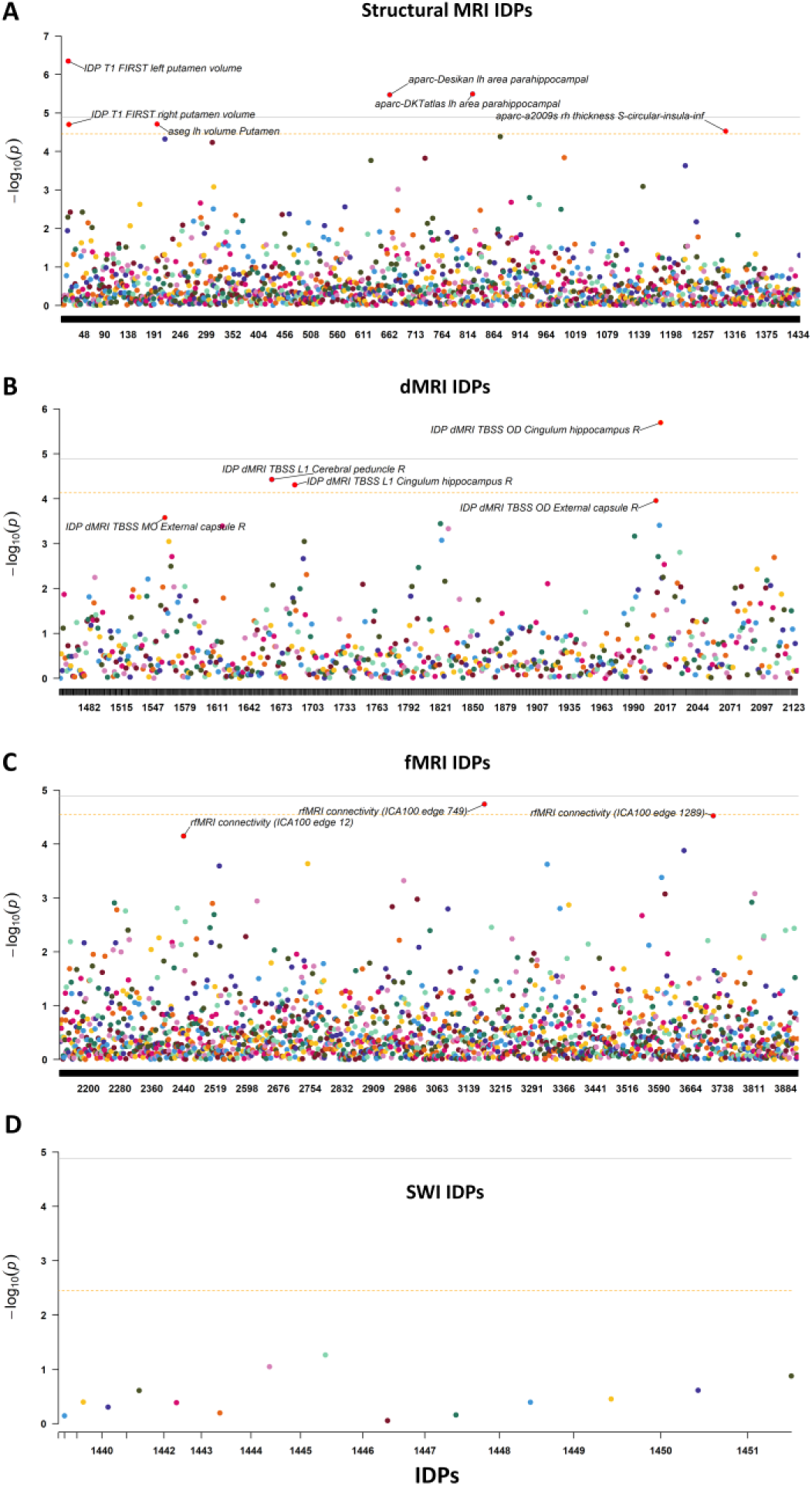
Causal associations between MS risk and brain IDPs in reverse MR analysis. (A) Causal associations between MS risk and 1433 structural MRI IDPs. (B) Causal associations between MS risk and 675 dMRI MRI IDPs. (C) Causal associations between MS risk and 1787 fMRI IDPs. (D) Causal associations between MS risk and 14 SWI IDPs. Causal effects were estimated using IVW method. Bonferroni-corrected *P* < 0.05 (uncorrected *P* < 1.28 x 10^-5^ [0.05/3909]) was considered statistically significant (Solid line). Bonferroni-corrected *P* < 0.05 within each category of IDPs was suggested to have potential significance (Dashed line). Abbreviations: IDP, Imaging-derived phenotype; MRI, Magnetic resonance imaging; SWI, Susceptibility-weighted imaging.

**Fig. 6.**
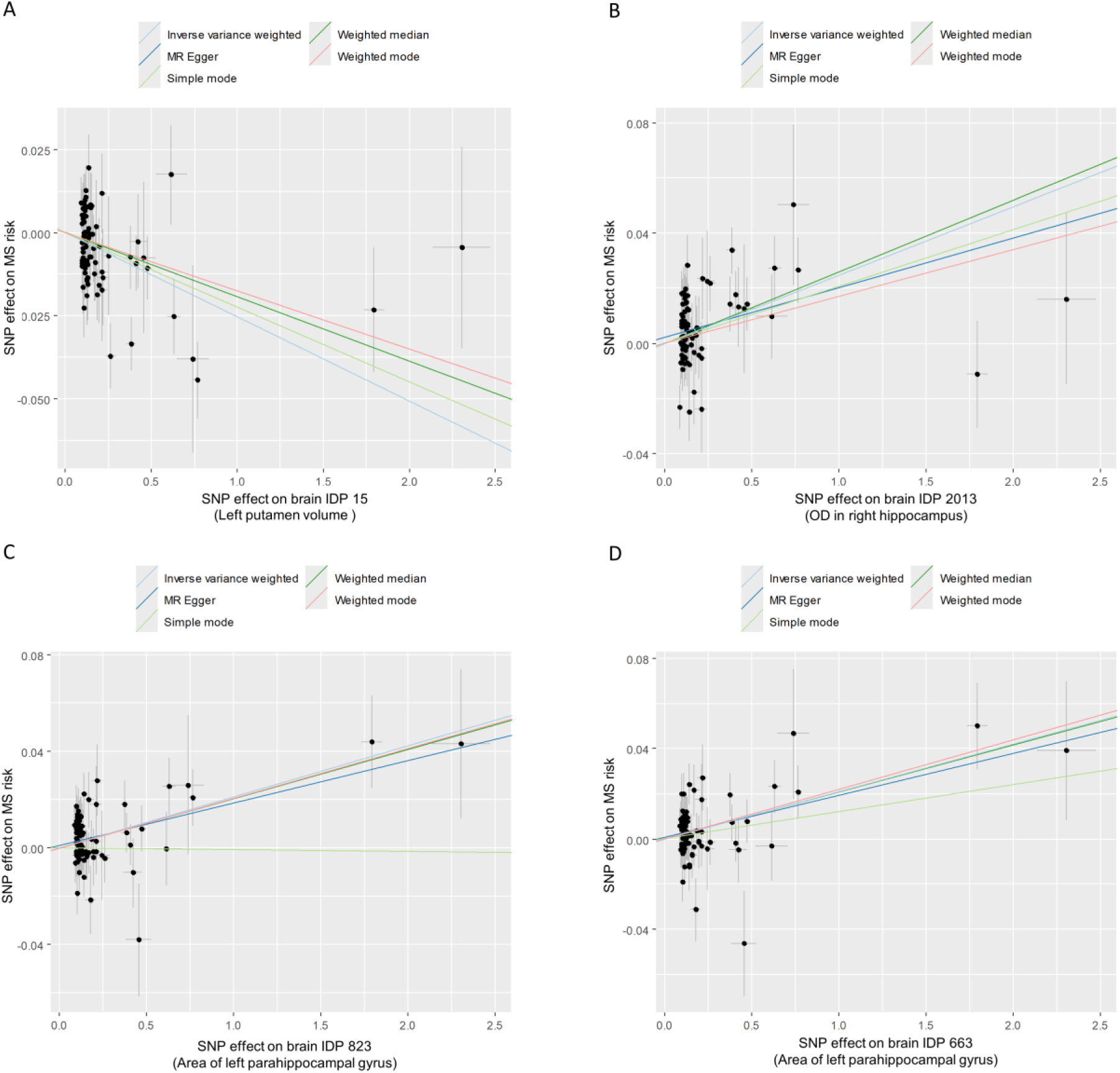
Putatively causal effects of MS risk on brain IDPs. (A-I) Causal effects of MS risk on 4 brain IDPs: IDP 15 (A, Left putamen volume), IDP 2013 (B, OD in right hippocampus), IDP 823 (C, Area of left parahippocampal gyrus), and IDP 663 (D, Area of left parahippocampal gyrus). Causal effects were estimated using five two-sample MR methods (MR-Egger, IVW, weighted median, weighted mode, and simple mode). Bonferroni-corrected *P* < 0.05 (uncorrected *P* < 1.28 x 10^-5^ [0.05/3909]) was considered statistically significant. Abbreviations: MS, Multiple sclerosis; IDP, Imaging-derived phenotype; SNP, Single nucleotide polymorphism; OD, Orientation dispersion index.

**Table 1.** The putative effects of NDDs on brain IDPs examined by MR analysis.

| Exposure<br>(NDDs) | UKB ID | Outcome<br>IDPs | OR (95%CI) <sup>a</sup> | *p*-value <sup>b</sup> | IV<br>number | Sensitivity<br>analysis <sup>c</sup> | Reverse effect <sup>d</sup> |
| --- | --- | --- | --- | --- | --- | --- | --- |
| AD |  |  |  |  |  |  |  |
| AD | 26578 | aseg rh intensity Amygdala | 1.06(1.04, 1.08) | 1.72E-09 | 27 | passed | No |
| AD | 25753 | rfMRI connectivity (ICA100 edge 681) | 1.06(1.04, 1.08) | 2.31E-08 | 27 | passed | No |
| AD | 26579 | aseg rh intensity Accumbens-area | 1.05(1.03, 1.07) | 3.33E-07 | 27 | passed | No |
| AD | 25891 | IDP T1 FAST ROIs R ventral striatum | 0.95(0.93, 0.97) | 4.68E-07 | 27 | passed | No |
| AD | 25754 | rfMRI amplitudes (ICA25 node 9) | 0.95(0.94, 0.97) | 3.58E-06 | 27 | passed | No |
| AD | 27032 | wg rh intensity-contrast inferiortemporal | 0.95(0.93, 0.97) | 4.54E-06 | 27 | passed | No |
| AD | 25754 | rfMRI amplitudes (ICA25 node 8) | 0.96(0.94, 0.97) | 6.24E-06 | 27 | passed | No |
| AD | 27039 | rfMRI amplitudes (ICA100 node 11) | 0.96(0.94, 0.98) | 8.01E-06 | 27 | passed | No |
| AD | 25755 | wg rh intensity-contrast parahippocampal | 0.96(0.94, 0.98) | 1.02E-05 | 27 | passed | No |
| MS |  |  |  |  |  |  |  |
| MS | 25015 | IDP T1 FIRST left putamen volume | 0.97(0.97, 0.98) | 4.47E-07 | 82 | passed | No |
| MS | 25428 | IDP dMRI TBSS OD Cingulum hippocampus R | 1.03(1.01, 1.04) | 2.02E-06 | 82 | passed | No |
| MS | 27156 | aparc-DKTatlas lh area parahippocampal | 1.02(1.01, 1.03) | 3.21E-06 | 82 | passed | No |
| MS | 26736 | aparc-Desikan lh area parahippocampal | 1.02(1.01, 1.03) | 3.37E-06 | 82 | passed | No |

**Abbreviations:** AD, Alzheimer's disease; NDDs: MR: Mendelian randomization; Neurogenerative diseases; IDP: Imaging-derived phenotypes; OR: Odds ratio; UKB: UK biobank; ICA: Independent component analysis; rfMRI: Resting-state functional magnetic resonance imaging; TBSS: Tract-Based Spatial Statistics; OD: orientation dispersion index.
<sup>a</sup>Inverse variance weighted (IVW) method was selected to be the major method to examine causal associations in MR analysis.
<sup>b</sup>Statistical significance was defined as Bonferroni-corrected $p < 0.05$ (uncorrected $p < 1.31E-05$ [0.05/3814]).
<sup>c</sup>The sensitivity analysis included heterogeneity test, LOO analysis, MR-PRESSO global test, and Pleiotropy test.
<sup>d</sup>The reverse MR analysis was conducted to examine the reverse effect.

## Discussion

Observational studies have reported significant relationships between brain IDPs and common NDDs; however, whether brain IDPs and NDDs are causally correlated remains poorly understood. In this study, we performed bidirectional two-sample MR analyses to systematically dissect the causal associations between 3909 IDPs and 4 NDDs. We demonstrated that AD was causally associated with 9 brain IDPs and MS was causally associated with 4 brain IDPs. We also identified one brain IDP was causally associated with increased risk of ALS. Therefore, our findings provided new insights into the bidirectional relationships between brain IDPs and NDDs.

In the forward MR analysis, we found most of brain IDPs were not causally associated with the risk of NDDs, which provided evidence that brain IDPs exerted weak effects on the prevalence of 4 NDDs. The major finding here is the causal association between greater volume of left cerebral white matter and increased risk of ALS. Consistently, Alexander *et al*. (2024) demonstrated that increased cerebral white matter volume was causally associated with higher risk of ALS^27^. However, according to previous literature, ALS patients tend to exhibit impairments of white matter integrity rather than increased white matter volume. For example, ALS patients exhibited significant white matter degeneration in frontal lobe, internal capsule, brainstem and hippocampal regions^28, 29, 30^. This discrepancy may be explained by the disease-specific white matter plasticity during childhood and adolescence^27^. White matter volume has been shown to be significantly increased through childhood and adolescence before slowly declining after the fourth decade^31^. White matter volume reduced with ageing and correlated with cognitive impiarment^32^, although mechanisms underlying age-related white matter loss are still not known. Whether the white matter volume in young preclinical ALS patients carrying disease-related genetic mutations differed from control individuals remained unknown. Interestingly, presymptomatic changes in brain volume have been observed in carriers of ALS/FTD-causing *MAPT* and *GRN* mutations in early adulthood, showing higher total intracranial volume in carriers compared with noncarriers^33^. However, it has been shown that ALS patients with hexanucleotide expansion in *C9orf72* showed significant bilateral degenerations in axonal structures of white matter along the corticospinal tracts and in fibers projecting to the frontal lobes^34^. Although these findings were divergent, they might suggest potential neurodevelopmental mechanisms in the initial stage of ALS occurrence. Therefore, the causal association between hemispheric white matter volume and ALS risk might be due to the differences in brain development during adolescence or childhood, which was shaped by genetic variations, age-dependent white matter plasticity, or systemic metabolic variables^27^. Overall, our findings provided evidence that higher brain white matter volume was a potential upstream element conferring elevated risk of ALS, thereby supporting a broad concept that brain IDPs influenced the risk of NDDs.

We identified reduced cortical thickness in left superior temporal pole was potentially associated with increased risk of AD, which was consistent with a recent MR study demonstrating that atrophy of the temporal pole was associated with higher AD risk^35^. In addition, MR analysis has shown that reduction in the surface area of left superior temporal gyrus was also associated with a higher risk of AD^36^. Previous studies have showed that cortical thickness in temporal pole was significantly reduced in AD patients^37, 38^ and correlated with apathy of the patients^39^. In addition, cortical thickness in temporal pole was also significantly associated with the severity of tau pathology measured with AV-1451-PET^40^. Nevertheless, whether it was the cause or the consequence of AD was an open question. Here, we provided evidence that it was atrophy of left superior medial temporal pole that putatively causes AD, but not AD led to the atrophy of left superior medial temporal pole (*P* > 0.05 in reverse MR analysis). Therefore, future prospective studies were encouraged to elucidate the role of left temporal pole in the prediction and diagnosis of AD.

AD patients exhibited widespread changes in structural, diffusion, and functional IDPs compared to control individuals^7, 41, 42, 43, 44^, nevertheless, whether genetically proxied AD causally shaped brain IDPs in patients remained poorly understood. Here, we revealed genetically determined risk of AD was causally associated with reduced volume of grey matter in right ventral striatum, which indicated that gray matter loss in right ventral striatum was a consequence of AD dementia^36^. Ventral Striatum played a key role in learning and memory^45^, as well as reward processing and motivated behavior^46, 47^. It was reported that ventral striatum exhibited stronger positive functional connectivity with the ventral caudate and medial orbitofrontal cortex, which were implicated in reward processing and motivation^48^. Selective loss of cholinergic neurons in the ventral striatum has been reported in AD patients^49^. In addition, amyloid-β (Aβ) has been shown to induce dopamine release in ventral striatum and decrease dopamine release in the dorsal striatum^50^. These findings might indicate that reward processing was impaired in AD. Indeed, impairment in reward processing has been revealed in AD mouse models^51, 52^. Additionally, abnormal reward behavior was also observed in typical AD patients^53^. The dysfunction of reward processing in AD patients might be associated with the significant disruptions of reward system in the brain, including ventral striatum^47^, as shown by our MR analysis. We found genetically proxied AD was also associated with higher mean intensity in amygdala^54, 55^ and accumbens area^56, 57^, which were both key components of reward system in brain. The atrophy of amygdala has been shown in patients with early AD^58^ and amygdala has been shown to play a key role in the propagation of neurofibrillary tangle pathology in AD^59^. In addition, amygdala atrophy was related to global cognitive functioning^60^ and the impairment of amygdala-frontal circuit has been shown to mediate the association between depressive symptoms and cognitive function in AD^61^. How mean intensity in amygdala changed in AD remained unknown, thereby deserving to be further explored in future studies. The impairment of nucleus accumbens has been shown to mediate reward processing dysfunction in AD. For example, loss of glycine receptors in the nucleus accumbens has been shown to induce the impairment of reward processing at an early stage of the disease^52^. In addition, age-dependent DAergic neuron loss in the ventral tegmental area has been shown to lead to lower DA outflow in the nucleus accumbens shell, which contributed to the dysfunction of reward processing^51^. In AD patients, lower within-network and between-network functional connectivity in reward networks (i.e., nucleus accumbens and orbitofrontal cortex) has been observed^62^. Taken together, genetically proxied AD was associated with altered brain IDPs involved in reward processing, thereby providing new insights into the neural mechanisms underlying reward dysfunction in AD patients.

We found genetically determined risk of AD was causally associated with reduced intensity-contrast in both right inferior temporal gyrus and right parahippocampal gyrus. Both right inferior temporal gyrus and right parahippocampal gyrus were core nodes within default mode network, which was significantly impaired in AD patients^63^. Patients with AD had significantly reduced gray matter volume^64^ and functional connectivity^65^ in the right inferior temporal gyrus compared to control individuals. In addition, white matter abnormalities and cortical thickness reduction in right parahippocampal gyrus have also been observed in AD patients^66, 67^. Therefore, AD was causally associated with impairments of core components belonging to default mode network.

We found genetically determined risk of MS was causally associated with reduced left putamen volume, which has been demonstrated by a recent observational study showing that MS patients exhibited lower volume in left putamen compared to control individuals^68^. MS was also associated with increased orientation dispersion index in right hippocampus, which was consistent with increased mean diffusivity in right hippocampus of MS patients^69^, indicating white matter abnormality in right hippocampus was a key feature of MS patients. We revealed MS was associated with increased area in left parahippocampal gyrus, which was consistent with a recent study showing a causal relationship between MS and area of parahippocampal gyrus^70^. Future studies were encouraged to validate these findings in MS patients.

Overall, this study used bidirectional MR method to investigate the causal associations between genetically determined brain IDPs and risk of 4 NDDs. The findings provided strong genetic evidence for possible causal links between brain IDPs and NDDs. This will help to develop better predictive imaging biomarkers and IDP-targeted interventions for NDDs at the brain-imaging level.

## Data Availability

Data availability
GWAS statistics of brain IDPs were collected from BIG40 web browser (https://open.win.ox.ac.uk/ukbiobank/big40).
Code availability
All software packages we used in the study are publicly available, and the download links are included in the Methods section. The Code was available from the corresponding author upon reasonable request.

## Acknowledgments

This work was supported by grants from National Natural Science Foundation of China (Grant No. 81873778, 82071415, and 82060213) and National Research Center for Translational Medicine at Shanghai, Ruijin Hospital, Shanghai Jiao Tong University School of Medicine (Grant No. NRCTM(SH)-2021-03).

## Supplementary Information

The online version contains supplementary material.

