## Supplemental file for "Causal Associations Between Imaging-derived Phenotypes and Risk of Alzheimer’s Disease and Other Neurodegenerative Disorders: A Mendelian Randomization Study"

### Supplementary materials

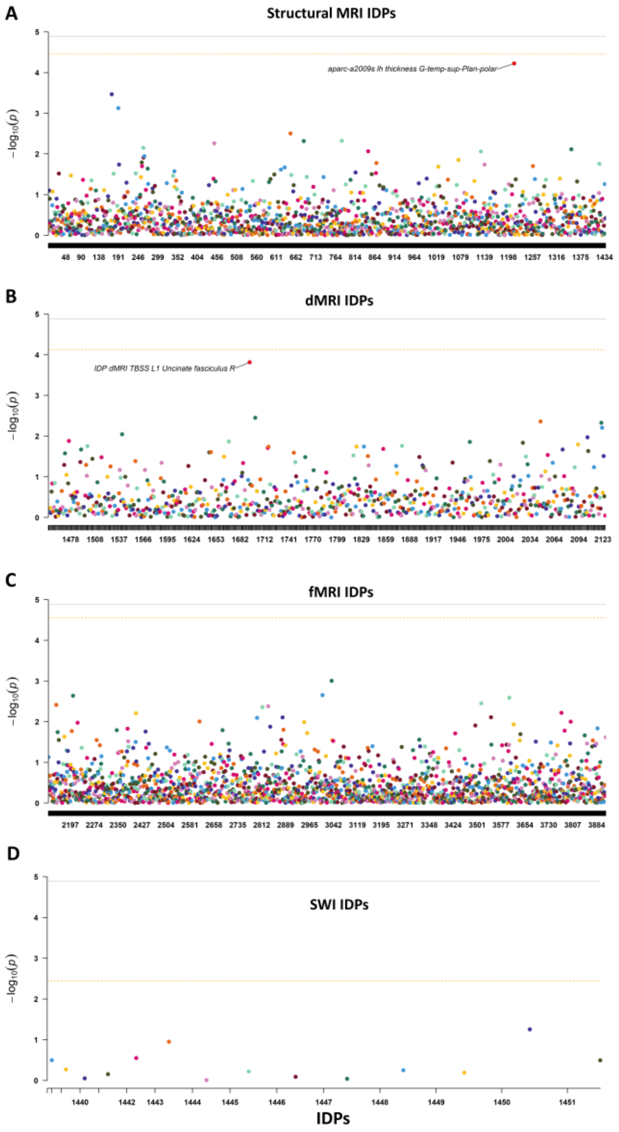


**Fig. S1 Causal associations between brain IDPs and AD risk in forward MR analysis.** (A) Causal associations between AD risk and 1433 structural MRI IDPs. (B) Causal associations between AD risk and 675 dMRI MRI IDPs. (C) Causal associations between AD risk and 1787 fMRI IDPs. (D) Causal associations between AD risk and 14 SWI IDPs. Causal effects were estimated using IVW method. Bonferroni-corrected $P < 0.05$ (uncorrected $P < 1.28 \times 10^{-5}$ [0.05/3909]) was considered statistically significant (Solid line). Bonferroni-corrected $P < 0.05$ within each category of IDPs was suggested to have potential significance (Dashed line). Abbreviations: IDP, Imaging-derived phenotype; MRI, Magnetic resonance imaging; SWI, Susceptibility-weighted imaging.

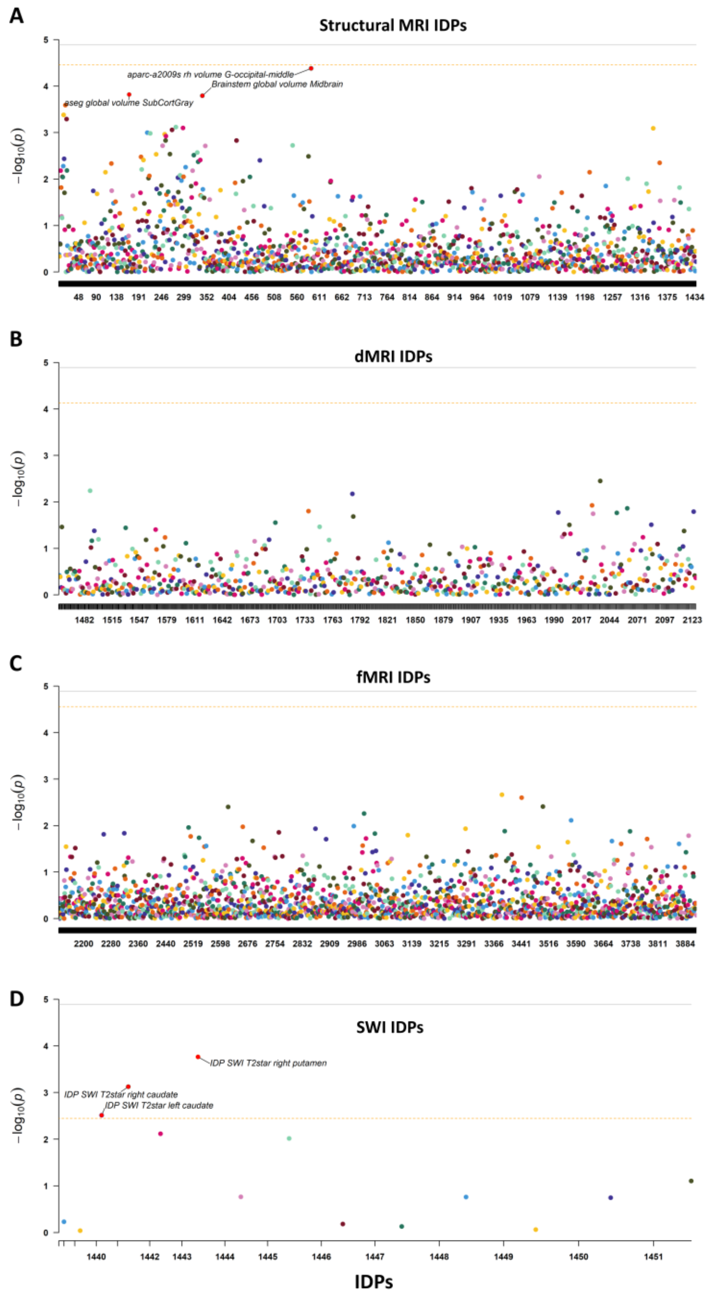


**Fig. S2 Causal associations between brain IDPs and PD risk in forward MR analysis.** (A) Causal associations between PD risk and 1433 structural MRI IDPs. (B) Causal associations between PD risk and 675 dMRI MRI IDPs. (C) Causal associations between PD risk and 1787 fMRI IDPs. (D) Causal associations between PD risk and 14 SWI IDPs. Causal effects were estimated using IVW method. Bonferroni-corrected $P$ < 0.05 (uncorrected $P$ < 1.28 x $10^{-5}$ [0.05/3909]) was considered statistically significant (Solid line). Bonferroni-corrected $P$ < 0.05 within each category of IDPs was suggested to have potential significance (Dashed line). Abbreviations: IDP, Imaging-derived phenotype; MRI, Magnetic resonance imaging; SWI, Susceptibility-weighted imaging.

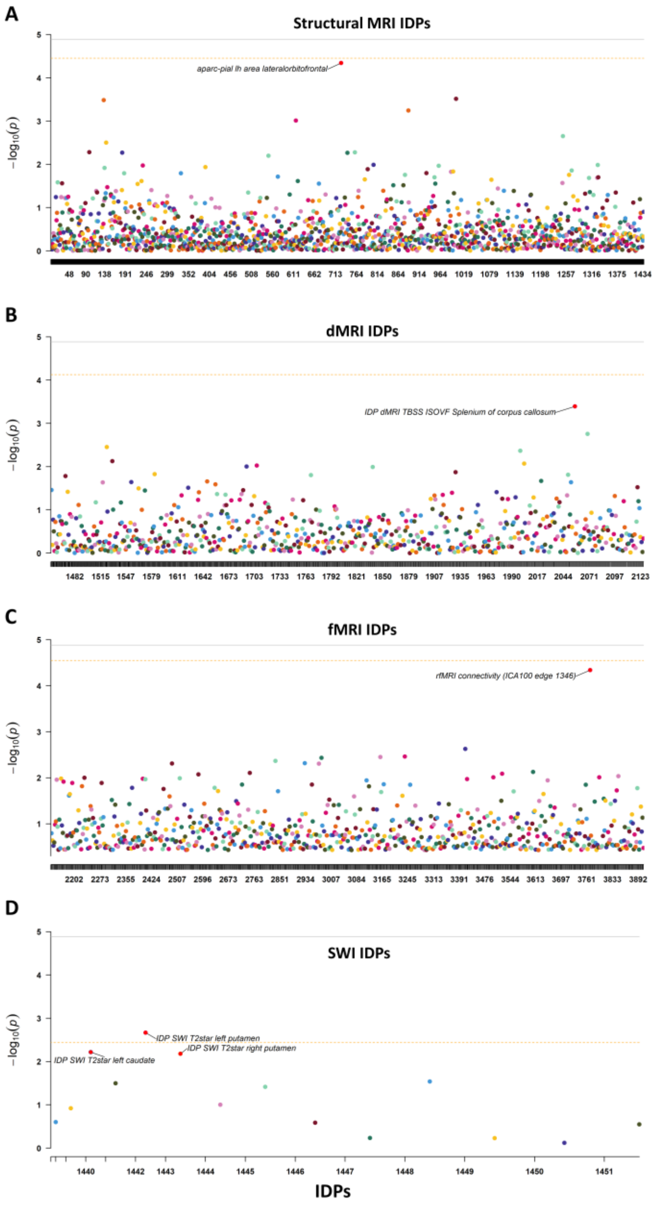


**Fig. S3 Causal associations between brain IDPs and MS risk in forward MR analysis.** (A) Causal associations between MS risk and 1433 structural MRI IDPs. (B) Causal associations between MS risk and 675 dMRI MRI IDPs. (C) Causal associations between MS risk and 1787 fMRI IDPs. (D) Causal associations between MS risk and 14 SWI IDPs. Causal effects were estimated using IVW method. Bonferroni-corrected $P$ < 0.05 (uncorrected $P < 1.28 \times 10^{-5}$ [0.05/3909]) was considered statistically significant (Solid line). Bonferroni-corrected $P$ < 0.05 within each category of IDPs was suggested to have potential significance (Dashed line). Abbreviations: IDP, Imaging-derived phenotype; MRI, Magnetic resonance imaging; SWI, Susceptibility-weighted imaging.

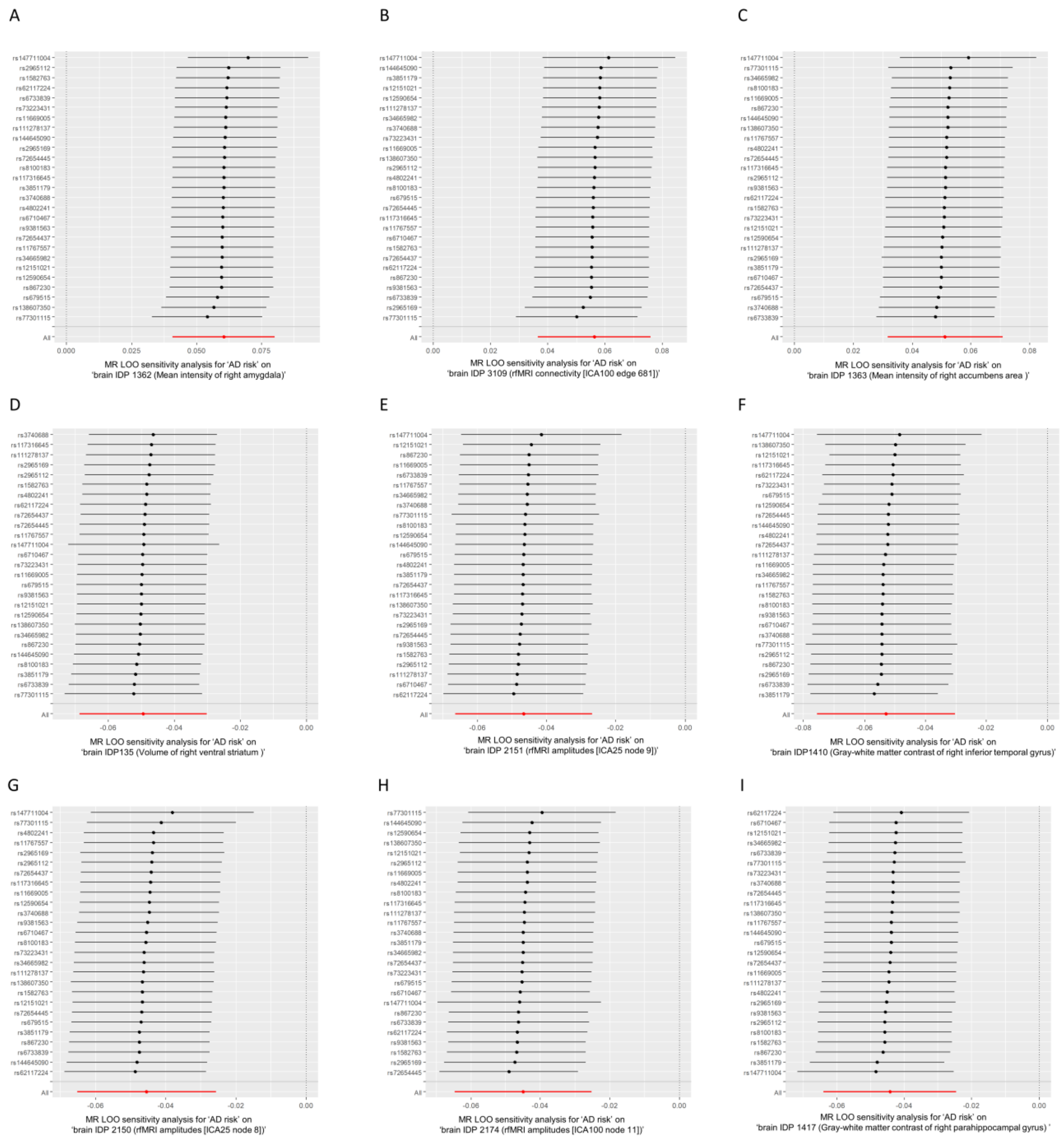

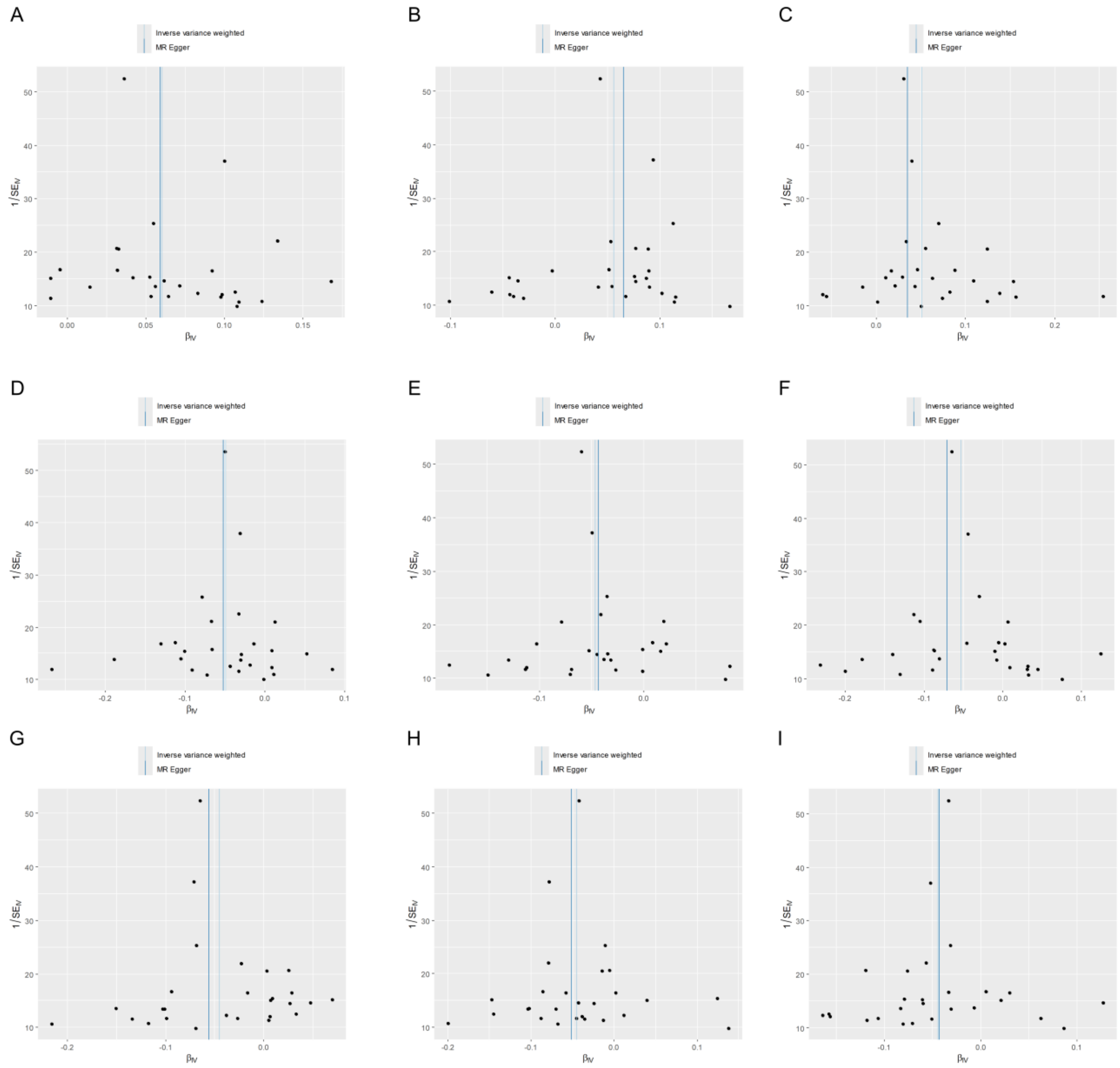


**Fig. S5 The funnel plot for significant MR causal estimates in Fig. 4.** (A-I) The funnel plot for MR results in Fig. 4A-I. Abbreviations: MR, Mendelian randomization.

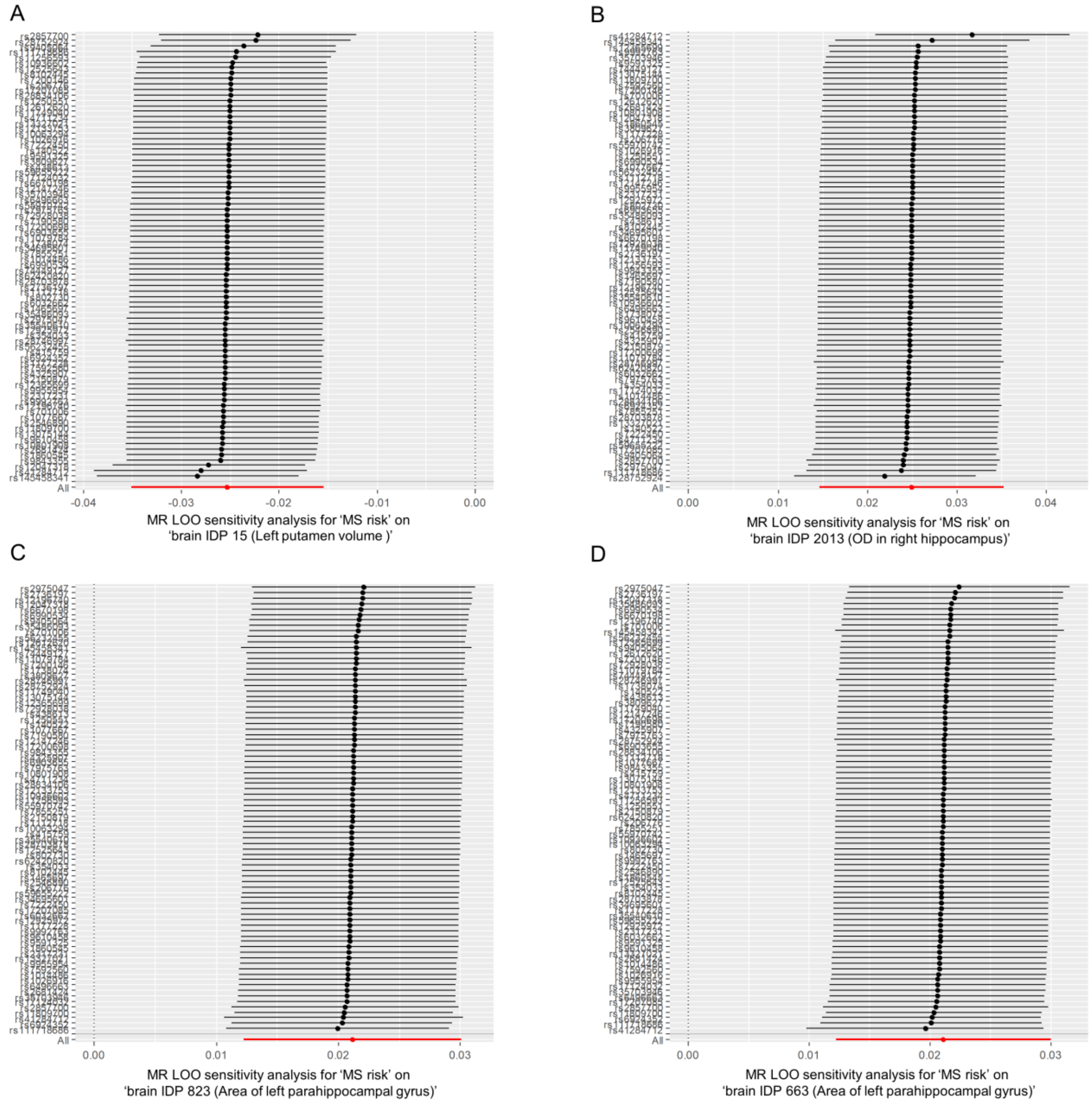


**Fig. S6 The LOO analysis for significant MR causal estimates in Fig. 6**. (A-D) The LOO analysis for significant MR causal estimates shown by Fig. 4A-D. Abbreviations: LOO, Leave-One-Out; MS, Multiple sclerosis; IDP, Imaging-derived phenotype; OD, Orientation dispersion index.

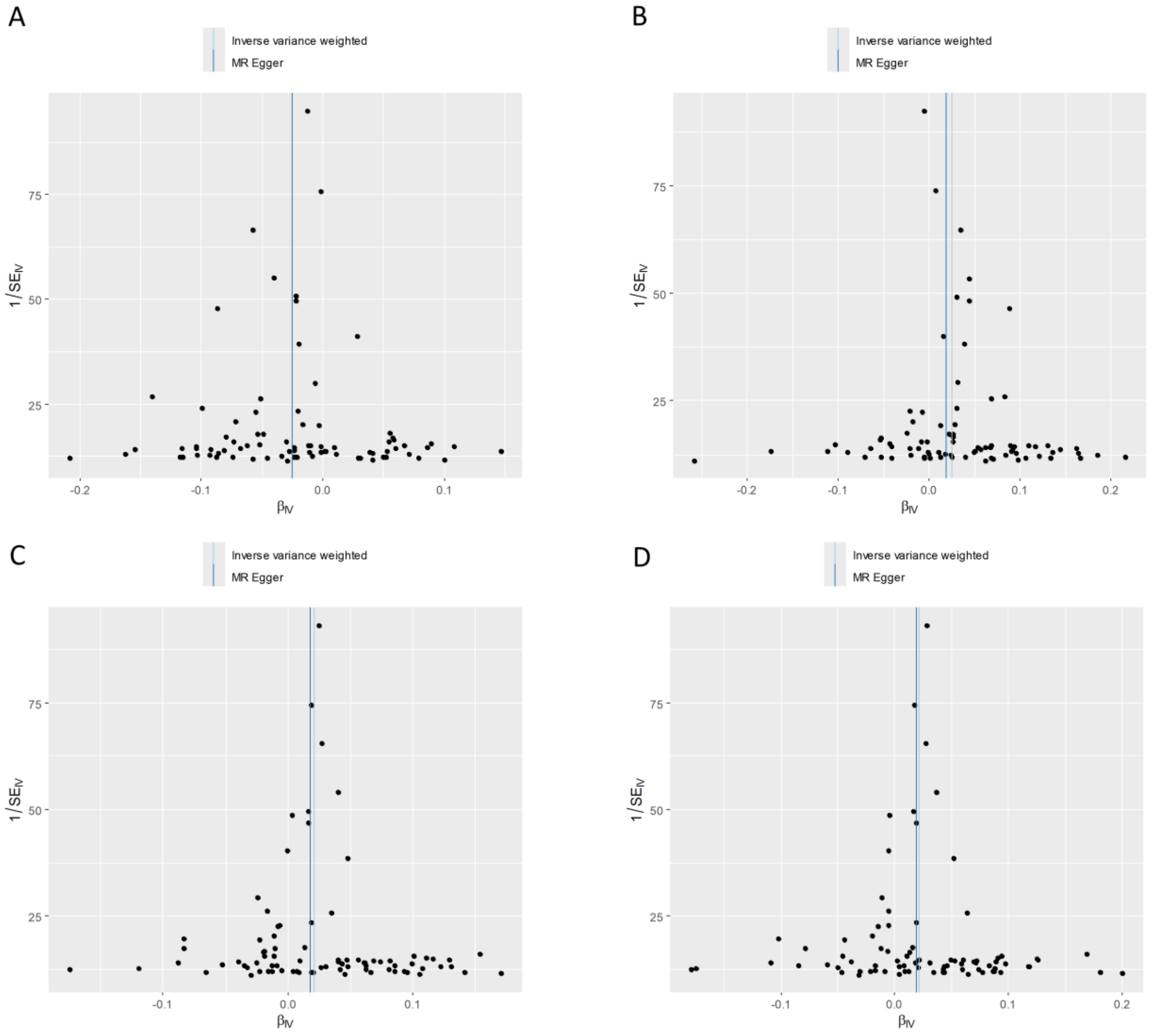


**Fig. S7 The funnel plot for significant MR causal estimates in Fig. 6.** (A-D) The funnel plot for MR results in Fig. 6A-D. Abbreviations: MR, Mendelian randomization.

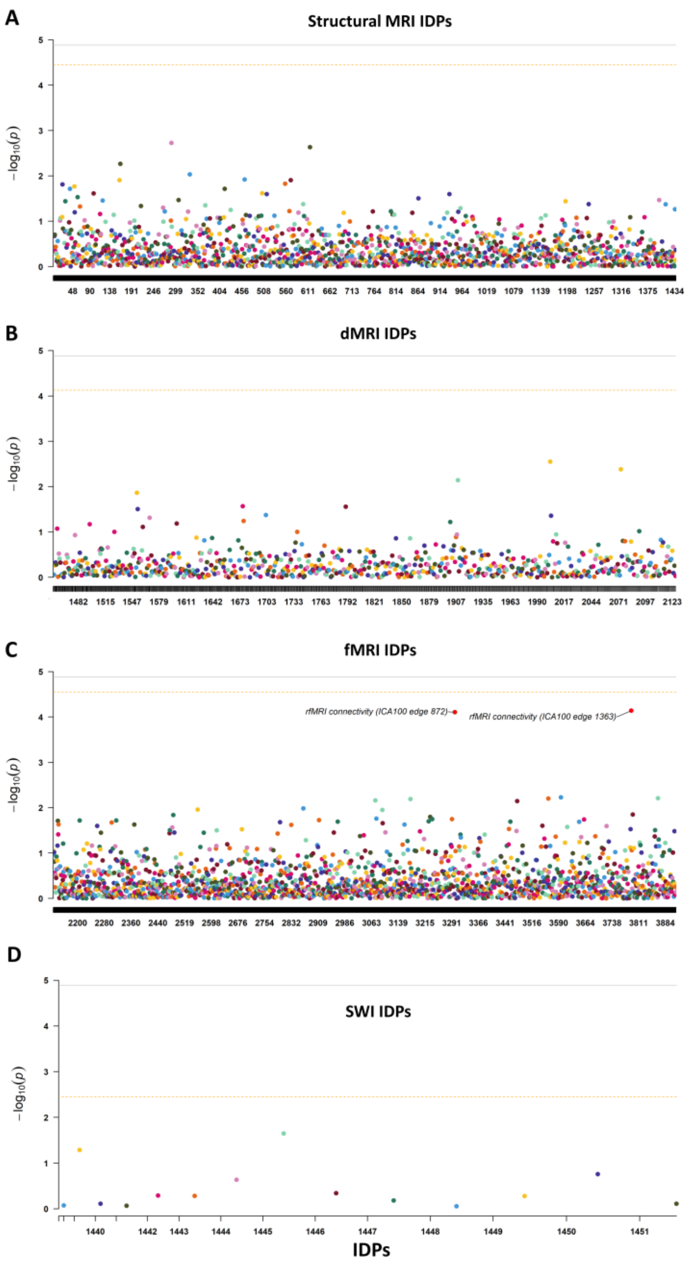


**Fig. S8 Causal associations between brain IDPs and PD risk in reverse MR analysis.** (A) Causal associations between PD risk and 1433 structural MRI IDPs. (B) Causal associations between PD risk and 675 dMRI MRI IDPs. (C) Causal associations between PD risk and 1787 fMRI IDPs. (D) Causal associations between PD risk and 14 SWI IDPs. Causal effects were estimated using IVW method. Bonferroni-corrected $P < 0.05$ (uncorrected $P < 1.28 \times 10^{-5}$ [0.05/3909]) was considered statistically significant (Solid line). Bonferroni-corrected $P < 0.05$ within each category of IDPs was suggested to have potential significance (Dashed line). Abbreviations: IDP, Imaging-derived phenotype; MRI, Magnetic resonance imaging; SWI, Susceptibility-weighted imaging.

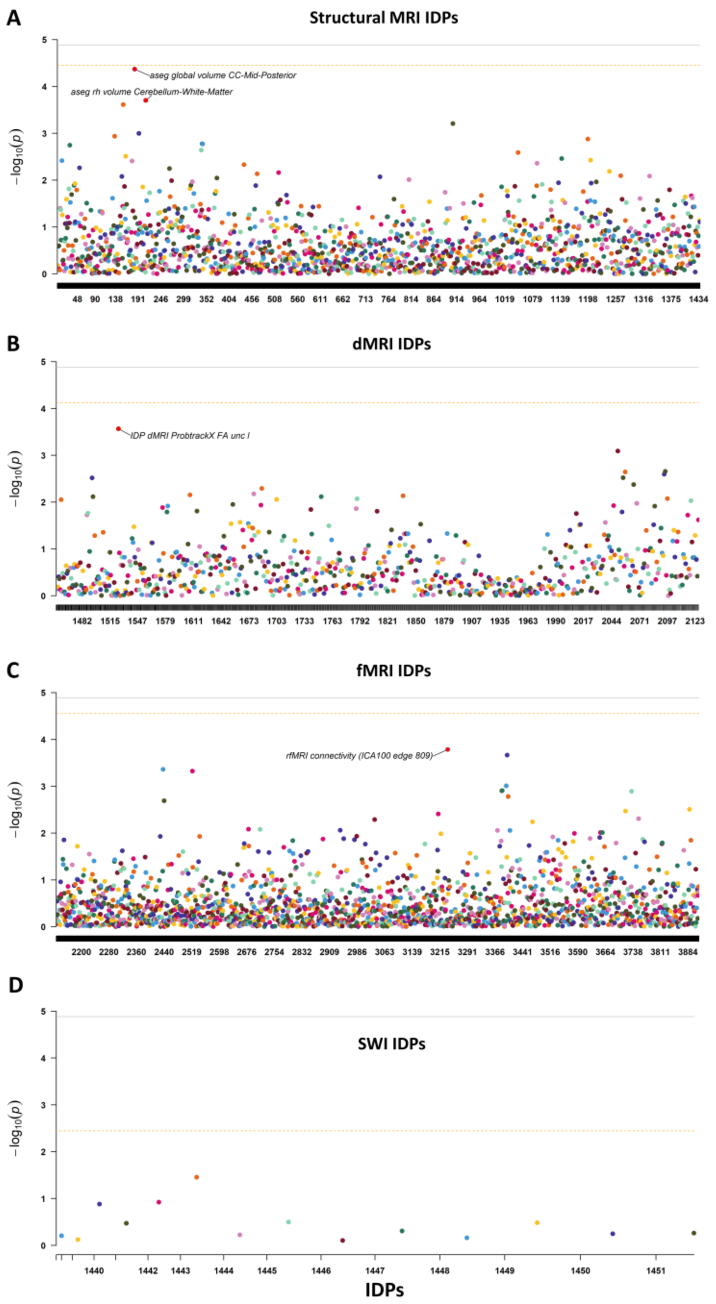


**Fig. S9 Causal associations between brain IDPs and MS risk in reverse MR analysis.** (A) Causal associations between MS risk and 1433 structural MRI IDPs. (B) Causal associations between MS risk and 675 dMRI MRI IDPs. (C) Causal associations between MS risk and 1787 fMRI IDPs. (D) Causal associations between MS risk and 14 SWI IDPs. Causal effects were estimated using IVW method. Bonferroni-corrected $P < 0.05$ (uncorrected $P < 1.28 \times 10^{-5}$ [0.05/3909]) was considered statistically significant (Solid line). Bonferroni-corrected $P < 0.05$ within each category of IDPs was suggested to have potential significance (Dashed line). Abbreviations: IDP, Imaging-derived phenotype; MRI, Magnetic resonance imaging; SWI, Susceptibility-weighted imaging.
